# Predicting up to 10 year breast cancer risk using longitudinal mammographic screening history

**DOI:** 10.1101/2023.06.28.23291994

**Authors:** Xin Wang, Tao Tan, Yuan Gao, Ruisheng Su, Tianyu Zhang, Luyi Han, Jonas Teuwen, Anna D’Angelo, Caroline A. Drukker, Marjanka K. Schmidt, Regina Beets-Tan, Nico Karssemeijer, Ritse Mann

## Abstract

Risk assessment of breast cancer (BC) seeks to enhance individualized screening and prevention strategies. BC risk informs healthy individuals of the short- and long-term likelihood of cancer development, also enabling detection of existing BC. Recent mammographic-based deep learning (DL) risk models outperform traditional risk factor-based models and achieve state-of-the-art (SOTA) at short-term risk prediction, but mainly use single-time exams, which seem to rely more on detecting existing lesions. We present a novel temporospatial and explainable deep learning risk model, the Multi-Time Point Breast Cancer Risk Model (MTP-BCR), which learns from longitudinal mammography data to identify subtle changes in breast tissue that may signal future malignancy. Utilizing a large in-house dataset of 171,168 screening mammograms from 42,792 consecutive exams involving 9,133 women, our model demonstrates a significant improvement in long-term (10-year) risk prediction with an area under the receiver operating characteristics (AUC) of 0.80, outperforming the traditional BCSC 10-year risk model and other SOTA methods at 5-year AUC in various screening cohorts. Furthermore, MTP-BCR provides unilateral breast-level predictions, achieving AUCs up to 0.81 and 0.77 for 5-year risk and 10-year risk assessments, respectively. The heatmaps derived from our model may help clinicians better understand the progression from normal tissue to cancerous growth, enhancing interpretability in breast cancer risk assessment.

**Teaser:** MTP-BCR model uses multi-time points mammograms and rich risk factors to predict 10-year breast cancer risk more accurately.

## Introduction

Breast cancer (BC) is one of the most common cancers in the world and is the cause of a large fraction of cancer-related mortality among women (*1, 2*). Studies have shown that age-based population-level BC screening programs, which aim to detect breast tumors at an early stage (*3*), reduces breast cancer specific mortality (*4–7*). However, the broad adoption of mammographic screening results also in high cost of imaging, false-positives and over-diagnoses, which explains the strong controversy of screening (*8, 9*). Therefore, “personalized” BC screening regimens are advocated, based on the individual women’s future risk of BC, which follows from demographic and genetic information, exposure to endogenous and exogenous risk factors, and also medical imaging (*10–12*). Current BC risk assessment models are designed to be sensitive to the high-risk population who could benefit from more aggressive screening and prevention. At the same time these models could advocate less frequent screening for the low-risk population to reduce the harm and cost of screening, however this is less common.

Based upon the timespan for breast cancer prediction, risk models can be divided into short- and long-term risk models. Short-term risk models can be used to guide physicians in selecting and adding supplemental screening modalities for women at the time of screening. Long-term risk prediction helps determine risk-based screening regimens and eligibility for preventive treatment (*13*). Many of the traditional risk models, such as Tyrer-Cuzick (*11*), CANRISK (*14*), National Cancer Institute Breast Cancer Risk Assessment Tool (BCRAT) (*12*), and Breast Cancer Surveillance Consortium (BCSC) (*15*) investigate primarily long-term risk estimates. The performances of these risk models remain modest in clinical practice and are not very sensitive to short/middle-term cancer risk variation due to the shortage of individual-specific risk adaptation, for example through the incorporation of detailed imaging findings beyond breast density only. With the recent boost in deep learning (DL) methods, some studies that combined large screening mammography datasets with detailed risk factors have shown considerable promise to help balance the harm-to-benefit ratios of BC screening (*7, 16–22*) and were even validated in clinical settings (*23*). For example, a recent study (*19*) developed a risk model, MIRAI, that achieved state-of-the-art (SOTA) performance in five-year BC risk prediction and outperformed the clinically adopted traditional models (*19, 23–25*).

However, most recent methods drive DL models to learn the risk output directly from an image or exam as single input without any historical reference (*7, 16, 19*). It’s like judging the motion trajectory of an object in a still frame of a video. We hypothesize that accurate estimation of the breast tissue changes may make the task of predicting long-term BC risk easier. In clinical practice, radiologists routinely compare mammography exams to identify developing abnormalities. Therefore, beyond learning risk features (e.g. breast density) from single-time point imaging, multi-time point learning may also be helpful in discovering the underlying dynamics of the risk pattern for BC development (*20, 26*). Moreover, due to the lack of a long-term longitudinal screening mammogram dataset, the potential of image-based DL methods for longer-term (e.g., 10 years) BC risk prediction has been less explored. Only one recent research investigated the long-term performance of an image-based short-term risk model (*27*).

Despite the development of promising risk models in BC screening programs, the interpretability of medical AI models is still difficult, whereas understanding the predicted outputs is essential for clinical acceptance. How to endow an existing risk model with explainability of the underlying reasoning remains the boundary to explore. Apart from being similar to what an actual radiologist does when searching for the sign of BC risk (*28, 29*), the AI models must reasonably show radiologists more details during inference (*30*) for clinical acceptance. However, most recent studies only aim to predict the patient-level risk and do not produce a location-specific risk. Improvement of these specific predictions and visualizations could not only improve the interpretability of the model and make it easier for physicians to understand the model’s decision-making but also inform doctors where to focus and then guide them when deciding on the most suitable targeted examinations and prevention strategies. An ideal risk model should therefore not only stratify high-risk groups but also focus the doctors attention to changing areas in the breast earlier.

We propose the Multi-Time-Points Breast Cancer Risk model (MTP-BCR), an end-to-end model that estimates the long-term future BC risk based on changes in breast tissue. Our contributions are as follows. First, our model leverages historical and current exams from a large in-house clinical mammogram dataset and obtains remarkable performance compared to other SOTA methods on patient-level BC risk prediction. Second, we explore and show that our image-based DL risk model outperforms clinical traditional BCSC risk models for long-term 10-year risk prediction. Third, we explore the unilateral breast level BC risk prediction and achieve similar performance to our risk models at the patient level. Fourth, we highlight suspicious areas in a longitudinal test dataset using the model’s heatmaps, which may illustrate the attention consistency of our model and improve its interpretability. Fifth, to demonstrate the robustness of our method in clinical settings, we perform a systematic subgroup analysis. The results imply that our model may improve upon traditional and other image-based DL risk models.

## Results

### Overview of algorithm

For investigating our hypothesis that the breast tissue changes can help in learning the tumor (including both invasive and ductal carcinoma in situ) development pattern better, multiple time points of examinations from the longitudinal screening mammograms are required. Like radiologists, who typically identify developing abnormalities by looking at changes in longitudinal exams, we propose a novel end-to-end multi-time point network, MTP-BCR, shown in Fig. 1A, leveraging longitudinal mammograms and medical records to capture the features related to increased BC risk. We aim to predict the risks on a patient-based level as well as for a single breast. Briefly, the proposed risk model first utilizes the multi-level (breast and patient level risk) and multi-task learning to extract static risk features from a single time point exam and prior medical records. Then the features of five historic exams, obtained before the current exam are selected (to mimic the practical use scenario) for learning the dynamic risk features using a multi-time point fusion model. This is combined with the risk factors of patients for predicting the future risk. It means that our end-to-end model uses current and historic screening mammography exams and existing medical records and then predicts the future 10-year BC risk. Architectural details, contents of the medical records, and risk factors are presented in the method part.

**Fig. 1.**
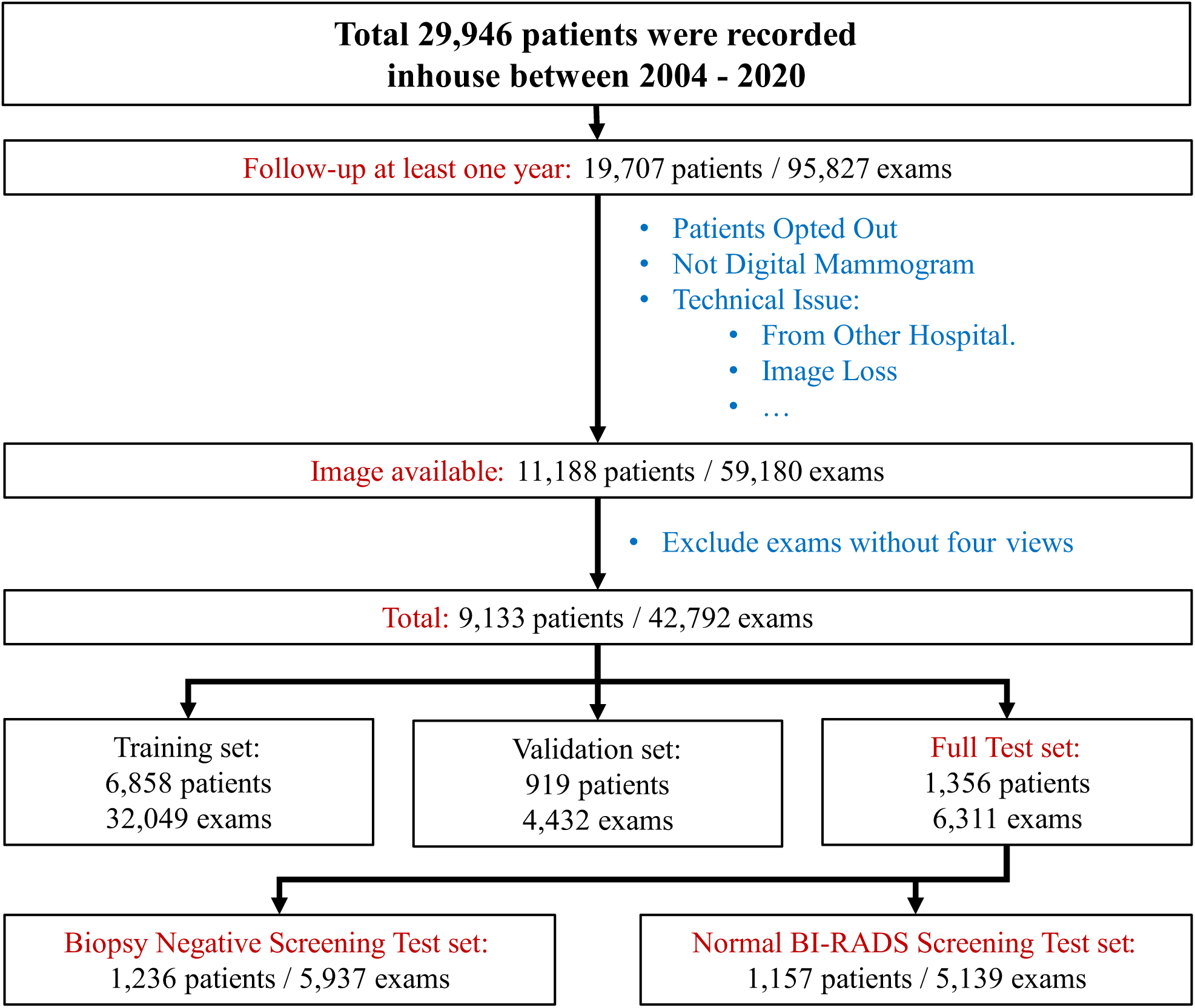
The flowchart of Inhouse mammogram dataset collection for 10-year risk prediction.

Risk calculation can be treated as a multi-class classification problem (*16, 19*), which is common in breast imaging, such as the classification of breast density (*31*), the Breast Imaging Reporting and Data System (BI-RADS (*32*)) score (*33*), the type of malignancy (*3*), and the BC molecular subtype (*34*). As shown in Fig. 1B, the risk of a patient getting BC within 5 or 10 years from the available data can be calculated as the cumulative sum of the probability from the first year up to the fifth or tenth year. Importantly, the prediction results of our model can guarantee that the risk is monotonically increasing and self-consistent. This avoids the situation, that can occur with separately trained models, where long-term risk may be lower than short-term risk. Moreover, this formulation also learns the inherent relationship between risks at different time points. In this study, the model is trained to predict the risk of BC at each of the 15 years and is validated by predicting BC within ten years. Therefore, our model can be easily extended for a longer than 15-year risk prediction when collecting enough longer-term follow-up data.

### Screening cohorts for risk modeling & compared candidates

Fig. 1 shows the flowchart of the Screening Cohort selection in the inhouse dataset. The dataset contains 42,792 exams of 9,133 patients, split into 32,049 exams / 6,858 patients, 4,432 exams / 919 patients, and 6,311 exams / 1,356 for the train set, validation set, and test set, respectively. The proposed model aims to handle multiple tasks, including cancer detection and future risk prediction. This would facilitate implementation in an actual clinical BC screening program, where not only focusing on the stratification of the high-risk population, short-term risk and determining whether women should be recalled is also essential. In fact, detection of existing malignancies, can be considered as an extremely short-term BC risk (*35*). Moreover, some of the model characteristics for the tasks of cancer detection and future risk prediction can be complementary (*19*).

To evaluate the model’s capability of longer term BC risk prediction, we use two standard screening test sets following the protocols of (*25*) and (*19*). The first one is named as the biopsy-negative screening group (5,937 exams / 1,236 patients), which includes mammography exams with BI-RADS 1 and 2 scores, or other BI-RADS scores but with benign biopsy results within 90 days from the screening date. The second test set is called the normal BI-RADS screening group (5,139 exams / 1,157 patients), which only consists of the exams scored as BI-RADS 0, 1 and 2. It aims to explore how the model performs on high-risk population stratification when radiologists deem the exams not suspicious.

The inhouse mammogram dataset is coupled to patient derived classical risk factors that can be used in the clinical Breast Cancer Surveillance Consortium version 2 (BCSC, https://tools.bcsc-scc.org/BC5yearRisk/). The distribution of clinical risk factors is shown in Supplemental Table S1. The BCSC model can estimate five-year, and ten-year BC risk based on risk factors but requires excluding patients following exclusion criteria (previous diagnosis of BC, younger than age 35 or older than age 74, or missing density estimates). Although studies have shown that image-based DL risk models outperform traditional risk models in 5-year risk assessments (*19, 24, 25*), the potential advantages of the former still need to be explored in longer-term 10-year risk assessments. Thus, we compare our model with not only the traditional BCSC 1-year and 5-year risk models but also with the BCSC predicted 10-year risk. To demonstrate the added value of the inclusion of patient based risk factors, we also build a similar multi-time point model without risk factors as MTP-BCR for comparison.

To demonstrate the risk prediction capacity, we compare it to the SOTA MIRAI (Massachusetts Institute of Technology, Boston, Massachusetts) model (*19*). MIRAI is a mammogram-based risk model that can predict 5-year risk at multiple time points and outperforms traditional models. Moreover, this model includes a pretrained risk factor predictor that allows missing risk factors. In this study, we also explore MIRAI’s ability of the longer-term 10-year risk estimate. We obtain the pretrained MIRAI model from their public GitHub (https://github.com/yala/Mirai). For a competitive comparison, we finetune the MIRAI model on the inhouse training dataset to alleviate the impact of domain shift.

We conduct hyperparameter search to finetune MIRAI and select the model with the best concordance index (C-index) on the validate set. Similar to the research (*19*), to investigate the performance of our method on BC detection, we also compare our model with the retrospective radiologist BI-RADS scores and the Globally Aware Multiple Instance (GMIC, New York University, New York) model (*29*). The GMIC is another recent SOTA DL model which focuses on detecting BC within three months, and some researches also show its potential for BC risk prediction (*19,25*). The pretrained GMIC model is obtained from the public GitHub repository (https://www.github.com/nyukat/gmic). For fairness, we re-implement the prepossessing from the raw DICOM format mammograms through their preset prepossessing pipeline and collect the ensembled predictions from the five pretrained models.

Note that, for full leverage of the mammogram examinations, we include all exams with at least one-year screening follow-up. To fairly compare five-year risk prediction with other SOTA methods and prove the contribution of our algorithm design of longitudinal input and multi-task learning, we also re-form the inhouse five-year risk dataset by excluding the examinations without at least five-year screening follow-up. Then we train our model from scratch on five-year risk prediction using the inhouse five-year risk dataset and we reperform all experiments (shown in the Supplemental Section).

### Risk prediction on full inhouse test dataset

All concordance index (C-index) and Area Under the Receiver Operating Characteristics (AUC) results on the inhouse test dataset (6,311 examinations / 869 positives within 5 years / 1,132 positives within 10 years) are summarized in Table 2.

**Table 1.**
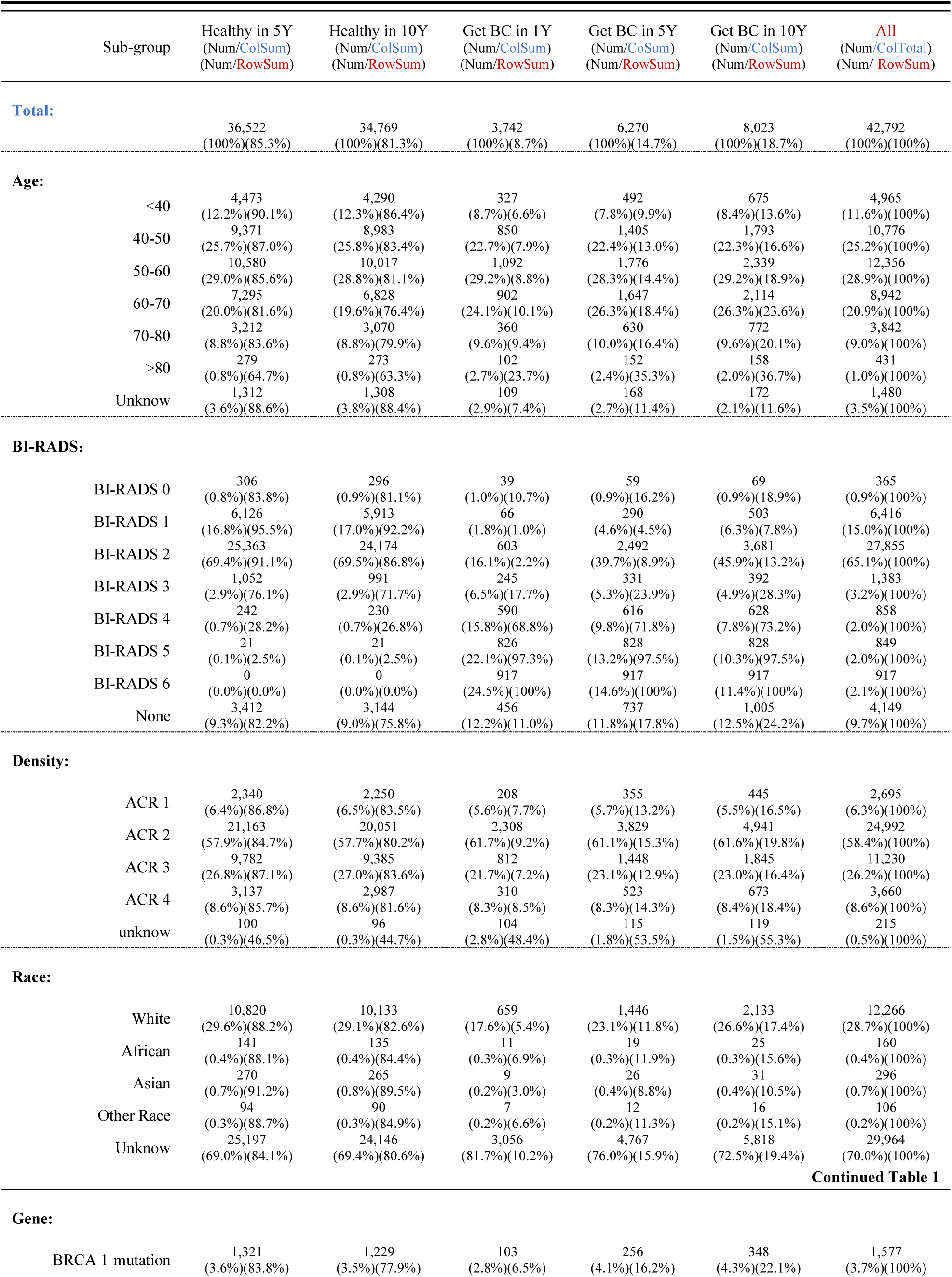
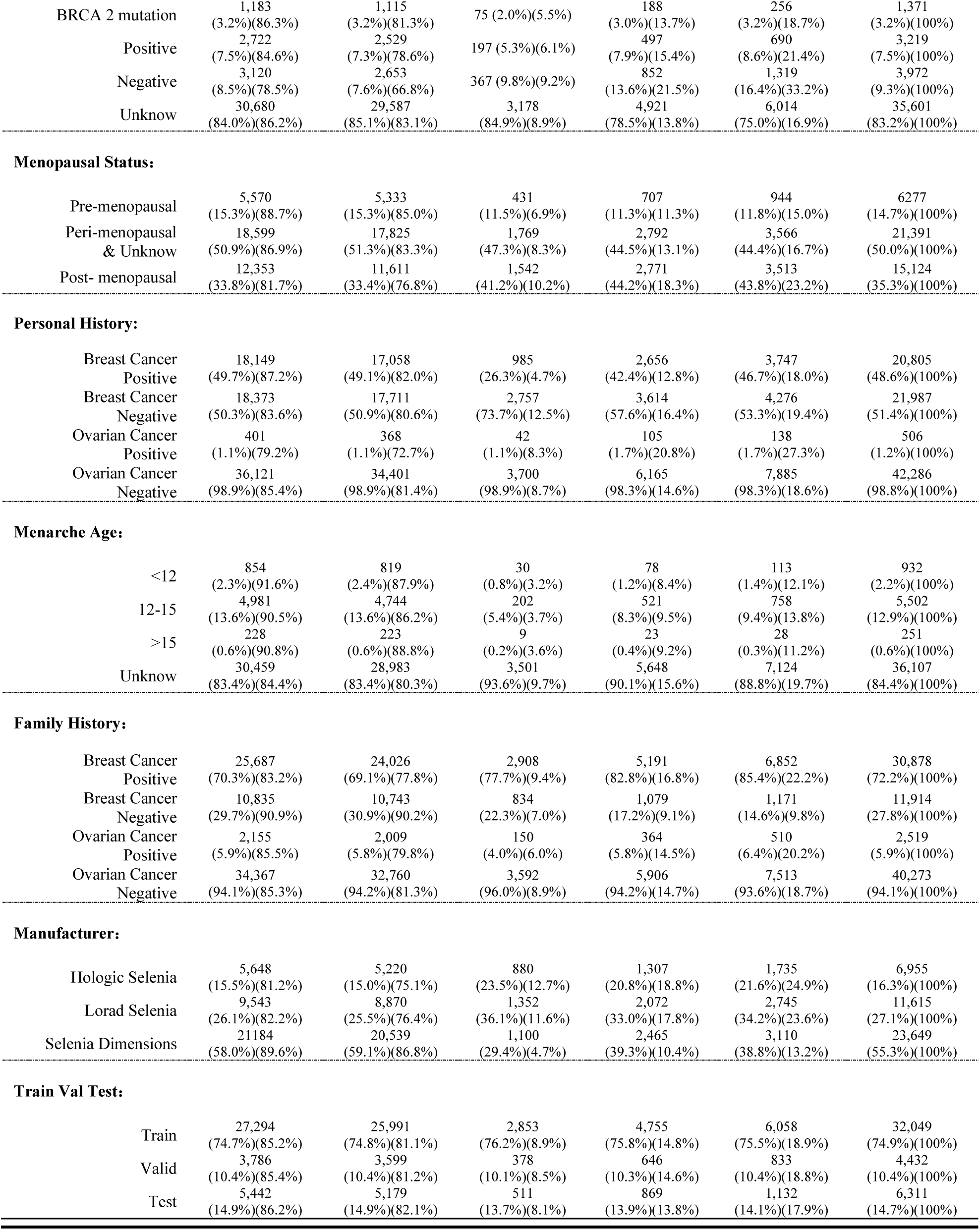
Detailed demographics for the Inhouse dataset. We categorize the number of mammography examinations into different demographic subgroups (rows) and different health subgroups (columns). The reported percentages are the number of examinations as a proportion of the total number of exams in the corresponding health conditions (Num/ColSum) and as a ratio of all exams in the same demographic sub-group (Num/RowSum).

**Table 2.** Comparison of 10-year risk predictions on full test set. C-index and AUC results are presented with 95% Confidence Interval. Note that results of the BCSC model are based on the part of the full inhouse test set, as they are out of the age range of 35-74 or with prior BC history. For a fair comparison, we also implemented the comparison experiments excluding the women that did not have scores from the BCSC model, Shown in Fig. 3. The black fonts represent the performance of the target tasks for which different methods were originally designed to. The gray fonts represent the AUC metric of 1- to 10-years BC risk for these methods we explored. Bold: *P* < 0.05, the AUCs of our methods are significantly higher than all other model for the same time horizon (except BCSC risk model).

| Use Risk Factors | BI-RADS | BCSC* | GMIC | MIRAI Test | MIRAI Finetune | MTP-BCR (Ours) Patient Level |  | MTP-BCR (Ours) Unilateral Breast Level |  |
| --- | --- | --- | --- | --- | --- | --- | --- | --- | --- |
|  | - | Yes | No | - | - | No | Yes | No | Yes |
| Full inhouse test set: 6,311 exams, 511 followed by cancer diagnosis within 1 years; 869 diagnosis within 5 years; 1,132 diagnosis within 10 years. |  |  |  |  |  |  |  |  |  |
| 5-Year | 0.71 | 0.63 | 0.7 | 0.67 | 0.75 | 0.78 | 0.84 | 0.78 | 0.82 |
| C-Index | (0.70-0.73) | (0.61-0.65) | (0.68-0.72) | (0.65-0.69) | (0.73-0.77) | (0.76-0.80) | (0.82-0.85) | (0.76-0.80) | (0.81-0.84) |
| 10-Year | 0.69 | 0.64 | 0.69 | 0.67 | 0.73 | 0.77 | 0.82 | 0.76 | 0.81 |
| C-Index | (0.67-0.70) | (0.61-0.66) | (0.67-0.70) | (0.65-0.68) | (0.72-0.75) | (0.75-0.78) | (0.81-0.84) | (0.75-0.78) | (0.79-0.82) |
| 1-Year | 0.83 | 0.62 | 0.74 | 0.70 | 0.84 | 0.87 | 0.91 | 0.87 | 0.89 |
| AUC | (0.81-0.85) | (0.58-0.64) | (0.72-0.77) | (0.68-0.73) | (0.81-0.86) | (0.85-0.89) | (0.89-0.92) | (0.85-0.89) | (0.87-0.91) |
| 2-Year | 0.78 | 0.62 | 0.72 | 0.69 | 0.80 | 0.83 | 0.88 | 0.83 | 0.87 |
| AUC | (0.76-0.80) | (0.59-0.65) | (0.70-0.75) | (0.66-0.71) | (0.78-0.82) | (0.81-0.85) | (0.86-0.89) | (0.81-0.85) | (0.85-0.88) |
| 3-Year | 0.74 | 0.63 | 0.72 | 0.69 | 0.78 | 0.80 | 0.86 | 0.80 | 0.84 |
| AUC | (0.72-0.76) | (0.61-0.66) | (0.69-0.74) | (0.67-0.72) | (0.76-0.80) | (0.79-0.82) | (0.84-0.87) | (0.79-0.82) | (0.83-0.86) |
| 4-Year | 0.72 | 0.64 | 0.71 | 0.69 | 0.76 | 0.79 | 0.84 | 0.79 | 0.82 |
| AUC | (0.70-0.73) | (0.61-0.67) | (0.69-0.73) | (0.67-0.71) | (0.74-0.78) | (0.77-0.80) | (0.82-0.85) | (0.77-0.81) | (0.81-0.84) |
| 5-Year | 0.70 | 0.65 | 0.70 | 0.70 | 0.74 | 0.77 | 0.82 | 0.77 | 0.81 |
| AUC | (0.68-0.71) | (0.62-0.68) | (0.68-0.72) | (0.68-0.72) | (0.72-0.76) | (0.75-0.79) | (0.81-0.84) | (0.75-0.79) | (0.79-0.82) |
| 6-Year | 0.68 | 0.66 | 0.70 | 0.70 | 0.73 | 0.77 | 0.82 | 0.76 | 0.80 |
| AUC | (0.67-0.70) | (0.63-0.69) | (0.68-0.72) | (0.68-0.72) | (0.71-0.75) | (0.75-0.79) | (0.80-0.83) | (0.75-0.78) | (0.78-0.81) |
| 7-Year | 0.67 | 0.67 | 0.70 | 0.70 | 0.73 | 0.76 | 0.81 | 0.76 | 0.79 |
| AUC | (0.66-0.69) | (0.65-0.70) | (0.68-0.72) | (0.68-0.72) | (0.71-0.75) | (0.74-0.78) | (0.80-0.83) | (0.74-0.77) | (0.77-0.80) |
| 8-Year | 0.66 | 0.69 | 0.69 | 0.69 | 0.72 | 0.76 | 0.80 | 0.75 | 0.78 |
| AUC | (0.65-0.68) | (0.66-0.71) | (0.67-0.71) | (0.67-0.71) | (0.70-0.74) | (0.74-0.78) | (0.79-0.82) | (0.74-0.77) | (0.76-0.79) |
| 9-Year | 0.65 | 0.70 | 0.69 | 0.68 | 0.71 | 0.76 | 0.80 | 0.75 | 0.77 |
| AUC | (0.64-0.67) | (0.67-0.73) | (0.67-0.71) | (0.66-0.70) | (0.69-0.73) | (0.74-0.78) | (0.78-0.81) | (0.74-0.77) | (0.75-0.78) |
| 10-Year | 0.65 | 0.71 | 0.69 | 0.67 | 0.71 | 0.77 | 0.80 | 0.76 | 0.77 |
| AUC | (0.63-0.66) | (0.68-0.74) | (0.66-0.71) | (0.65-0.70) | (0.68-0.73) | (0.75-0.79) | (0.78-0.82) | (0.74-0.77) | (0.75-0.78) |

#### Our method’s performance

The performances of the MTP-BCR model with risk factors and without risk factors at patient level risk prediction (as shown in Table 2, and ROCs are shown in Fig. S3) obtained 10-year C-indices of 0.82 (95% CI, 0.81-0.84) and 0.77 (95% CI, 0.75-0.78), with AUCs of 0.91 (95% CI, 0.89-0.92) and 0.87 (95% CI, 0.85-0.89) at easiest 1-year risk prediction and AUCs at the most difficult 10-year risk prediction of 0.80 (95% CI, 0.78-0.82) and 0.77 (95% CI, 0.75-0.79), respectively. The AUCs results of 1- to 10-year risk prediction show that the performances of the MTP-BCR with risk factors are significantly higher than those of MTP-BCR without risk factors (All *P* values < 0.05). We also evaluate the unilateral breast level cancer prediction of MTP-BCR models both with risk factors and without risk factors, as shown in Fig. S3. Similar performances of patient-level risk prediction are obtained, with the 10-year C-indices of 0.81 (95% CI, 0.79-0.82) and 0.76 (95% CI, 0.75-0.78) for our methods with and without risk factors, and 5-year C-indices of 0.82 (95% CI, 0.81-0.84) and 0.78 (95% CI, 0.76-0.80). The AUCs of unilateral breast-level cancer prediction without risk factors ranged from 0.76 to 0.87. When incorporating patient derived risk factors, the AUCs ranged from 0.77 to 0.89. Therefore, our MTP-BCR risk model can also accurately predict the risk of development of BC in a unilateral breast, and using risk factor information can further improve the performance of 1- to 10-year risk.

Moreover, we perform two ablation studies to choose the best design of the MTP-BCR model (Table S4 and S5). The first one is to investigate whether our multi-task and multi-level, and multi-time point learning strategies can improve the ability to extract risk-related features. The C-indices and AUCs show that the model with multi-task, multi-level, and multi-time-point learning is better than others alone. Using risk factors can further improve the performance of the risk models. Besides, the model achieves the best performance when using five time-point historic mammogram references.

#### Comparing with other methods (except BCSC)

The 1-year AUCs of Radiologists BI-RADS assessments and the BC detection method GMIC are 0.83 (95% CI, 0.81-0.85) and 0.74 (95% CI, 0.72-0.77) respectively, which are significantly lower than both MTP-BCR risk models (*P* < 0.001). Therefore our MTP-BCR risk model outperforms radiologists and the SOTA BC detection model for BC detection and extremely short-term risk prediction, even when only using mammograms. Comparing to the SOTA DL-based MIRAI model, the 5-year C-indices of the MTP-BCR models (with/without risk factor) are 0.82 (95% CI, 0.81-0.84) and 0.77 (95% CI, 0.75-0.78) versus 0.73 (95% CI, 0.72-0.75). The AUC results show that both the MTP-BCR models significantly outperform the MIRAI model at all time points from 1-year to 5-year risk (All *P* values < 0.01). Also for 10-year risk prediction, the MTP-BCR methods still have a significant advantage (All *P* values < 0.05).

#### Comparing with BCSC risk model

Note that our study also include patients who are not eligible for risk calculation by the BCSC model as they are either out of the required age range of 35-74 or had a prior BC history. For this comparison, we conduct the experiments excluding the women that did not have scores from the BCSC model (Table S1). The AUC curves of all methods are shown in Fig. 3A. The AUC results show that both the MTP-BCR models significantly outperform the 1-year, 5-year, and 10-year BCSC risk models (All *P* values < 0.001). The latter obtains AUCs of 0.62 (95% CI, 0.58-0.64), 0.65 (95% CI, 0.62-0.68), and 0.71 (95% CI, 0.68-0.74), respectively. The 5-year and 10-year C-indices of BCSC models are 0.63 (95% CI, 0.61-0.65) and 0.64 (95% CI, 0.61-0.66), compared to C-indices of 0.91 (95% CI, 0.90-0.92) and 0.90 (95% CI, 0.88-0.91) by our MTP-BCR risk model with risk factors. Moreover, we also compare our risk models with the other methods in this specific population. The AUC results show that both the MTPBCR models significantly outperform all other models for 1-year to 10-year risk prediction (All *P* values < 0.001).

**Fig. 2.**
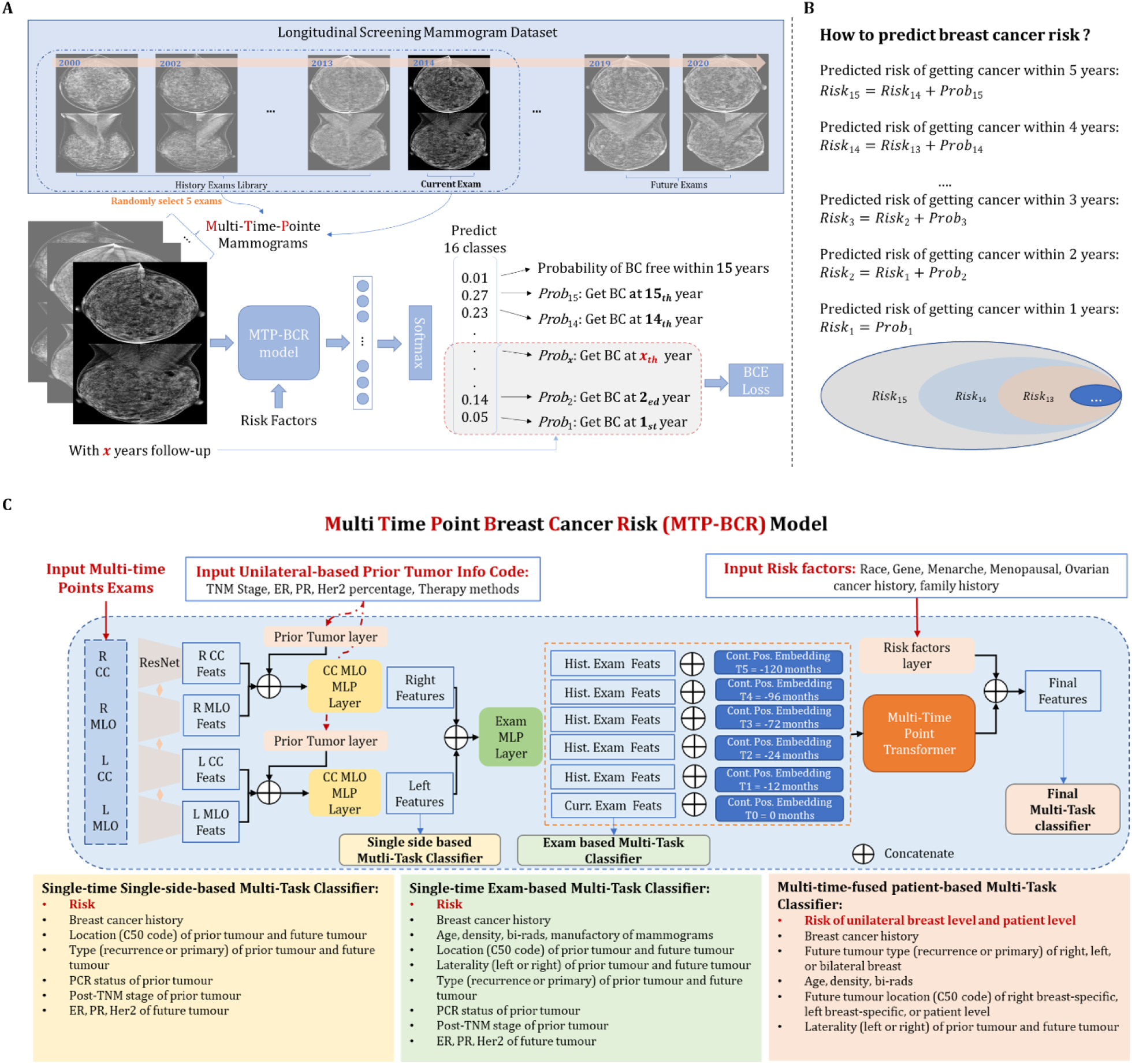
Schematic description of Multi-Time-Point Breast Cancer Risk (MTP-BCR) model. A. Overview of selecting the multi-time points mammograms for training the MTP-BCR model. B. how to calculate the BC risk C. The details of the MTP-BCR model

**Fig. 3.**
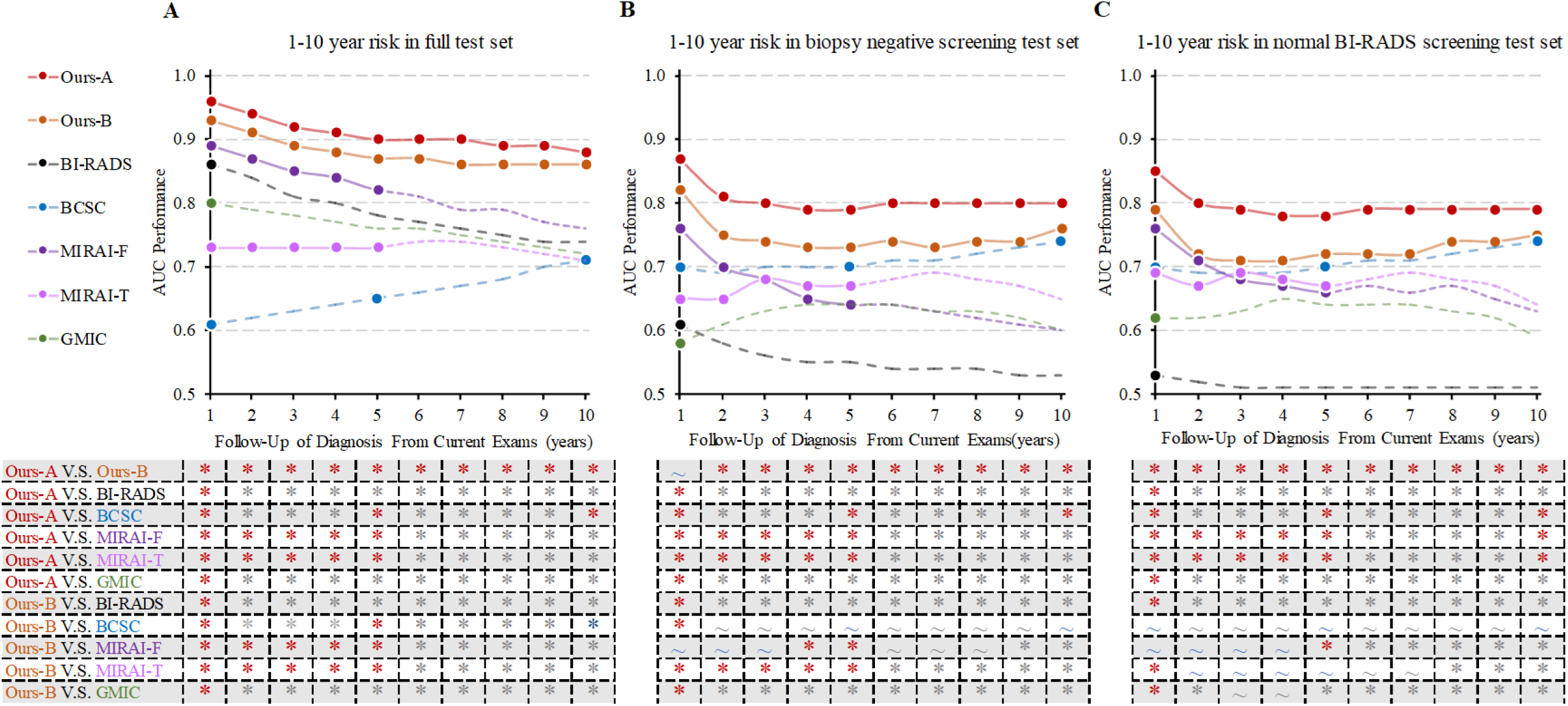
Cumulative risk at multiple time points (only for women who are completely scored across the BCSC model). Results on the full test set (A), biopsy negative screening test set (B), and normal BI-RADS screening test set (C) are shown in the left, middle, and right, respectively. Ours A: MTP-BCR with risk factors; Ours B: MTP-BCR without risk factors. The color dots represent the performance of the target tasks for which different methods are originally designed to. For instance, the MIRAI risk model is designed to predict 5-year BC risk at multiple time points, GMIC and BI-RADS focus on detecting BC within three months, and the BCSC risk model is used to predict 1-, 5-, and 10-year BC risk. Our methods are designed to predict 10-year BC risk at multiple time points. The dashed curves with different colors represent the AUC metric of 1- to 10-years BC risk for these methods we explored. Asterisk (*) means that there is a significant difference (DeLong’s test, P < 0.05) in two AUCs corresponding to the methods, whereas tilde (∼) indicates that there is no significant difference (DeLong’s test, P > 0.05) in AUCs of the two methods.

#### Performing experiments of 5-year risk prediction

Similar results are obtained for reperformed experiments on the formed 5-year risk prediction dataset (Table S6 and S7). Both the MTP-BCR models with and without risk factors are significantly better than other methods. Moreover, we also perform two same ablation studies on the inhouse 5-year risk dataset (Table S12 and S13). Similar results to those of 10-year risk ablation experiments also demonstrate the stability of our learning strategy.

### Risk prediction in a healthy screening population

#### In biopsy negative screening population

we evaluate all methods using the inhouse negative biopsy screening test set including 5,937 examinations / 495 positives within 5 years / 758 positives within 10 years, as shown in Table 3. In this test set, results show that our MTP-BCR with risk factors holds superiority in C-index and AUC metrics. Extremely short-term (1 year) risk prediction can be equivalent to interval cancer detection. In this task, the AUC of our model without risk factors is 0.70 (95% CI, 0.65-0.74). The AUC increases to 0.77 (95% CI, 0.73-0.81) when using risk factors, significantly higher than that from BI-RADS scores with 0.61 (95% CI, 0.57-0.65), the BC detection method GMIC with 0.59 (95% CI, 0.54-0.65), and also the finetuned MIRAI with 0.65 (95% CI, 0.60-0.70). In Fig. 3B and Table S2, the clinical BCSC 1-year risk model obtains an AUC of 0.70 (95% CI, 0.63-0.76), which is significantly lower than the AUCs of our MTP-BCR models (with risk factor: 0.87 / without risk factor: 0.82). For the long-term 5-year risk prediction, our model reaches a C-index of 0.65 (95% CI, 0.63-0.68) without risk factors and a C-index of 0.74 (95% CI, 0.72-0.76) with risk factors, versus 0.64 (95% CI, 0.62-0.66) for the finetuned MIRAI model (Table 3). When comparing with the clinical BCSC 5-year risk model (as shown in Table S1), our MTP-BCR obtains a C-index of 0.79 (95% CI, 0.76-0.82) versus 0.69 (95% CI, 0.65-0.72). For the longer-term 10-year risk estimate, our only image based MTP-BCR model has a similar performance to 10-year risk BCSC model according by the 10-year C-index. But the results indicate that the MTP-BCR with risk factors model still significantly outperforms all other models by AUCs at each time point (All *P* values < 0.05).

**Table 3.** Comparison of 10-year risk predictions on Screening Test sets. C-index and AUC results are presented with 95% Confidence Interval. Note that results of the BCSC model are based on part of the test sets-as they are out of the age range of 35-74 or with prior BC history. For fair comparison we also impended the comparison experiments excluding the women that did not score across the BCSC model Show as Fig. 3. The black fonts represent the performance of the target tasks for which different methods were originally designed to. The gray fonts represent the AUC metric of 1- to 10-years BC risk for these methods we explored. Bold: *P* < 0.05, the AUCs of our methods are significantly higher than all other model for the same time horizon (except BCSC risk model).

| Use Risk Factors | BI-RADS | BCSC * | GMIC | MIRAI Test | MIRAI Finetune | MTP-BCR (Ours) Patient Level |  | MTP-BCR (Ours) Unilateral Breast Level |  |
| --- | --- | --- | --- | --- | --- | --- | --- | --- | --- |
|  | - | Yes | No | - | - | No | Yes | No | Yes |
| Inhouse Biopsy Negative Screening Test set: 5,937 exams, 137 followed by cancer diagnosis within 1 years; 495 diagnosis within 5 years; 758 diagnosis within 10 years. |  |  |  |  |  |  |  |  |  |
| 5-Year | 0.54 | 0.69 | 0.61 | 0.63 | 0.64 | 0.65 | 0.74 | 0.65 | 0.73 |
| C-index | (0.53-0.56) | (0.65-0.72) | (0.59-0.64) | (0.60-0.65) | (0.62-0.66) | (0.63-0.68) | (0.72-0.76) | (0.63-0.67) | (0.71-0.75) |
| 10-Year | 0.53 | 0.69 | 0.61 | 0.62 | 0.64 | 0.65 | 0.74 | 0.65 | 0.72 |
| C-index | (0.52-0.55) | (0.66-0.72) | (0.59-0.63) | (0.60-0.64) | (0.62-0.66) | (0.63-0.67) | (0.72-0.76) | (0.63-0.67) | (0.71-0.74) |
| 1-Year | 0.61 | 0.70 | 0.59 | 0.63 | 0.65 | 0.70 | **0.77** | 0.68 | 0.76 |
| AUC | (0.57-0.65) | (0.63-0.76) | (0.54-0.65) | (0.58-0.68) | (0.60-0.70) | (0.65-0.74) | **(0.73-0.81)** | (0.62-0.73) | (0.71-0.81) |
| 2-Year | 0.58 | 0.69 | 0.61 | 0.62 | 0.64 | 0.67 | **0.75** | 0.66 | 0.75 |
| AUC | (0.55-0.60) | (0.64-0.75) | (0.57-0.65) | (0.58-0.66) | (0.60-0.67) | (0.63-0.70) | **(0.71-0.78)** | (0.62-0.70) | (0.72-0.79) |
| 3-Year | 0.55 | 0.70 | 0.62 | 0.65 | 0.65 | 0.66 | **0.75** | 0.66 | 0.74 |
| AUC | (0.54-0.58) | (0.65-0.74) | (0.59-0.65) | (0.62-0.68) | (0.62-0.68) | (0.63-0.69) | **(0.72-0.77)** | (0.63-0.69) | (0.71-0.77) |
| 4-Year | 0.54 | 0.70 | 0.63 | 0.64 | 0.65 | 0.66 | **0.74** | 0.66 | 0.72 |
| AUC | (0.53-0.56) | (0.65-0.74) | (0.60-0.65) | (0.62-0.67) | (0.62-0.68) | (0.63-0.68) | **(0.71-0.76)** | (0.63-0.68) | (0.70-0.75) |
| 5-Year | 0.54 | 0.70 | 0.62 | 0.65 | 0.66 | 0.66 | **0.73** | 0.66 | 0.72 |
| AUC | (0.52-0.55) | (0.65-0.74) | (0.60-0.65) | (0.63-0.68) | (0.63-0.68) | (0.63-0.68) | **(0.71-0.76)** | (0.63-0.68) | (0.69-0.74) |
| 6-Year | 0.53 | 0.71 | 0.63 | 0.65 | 0.67 | 0.66 | **0.74** | 0.66 | 0.72 |
| AUC | (0.52-0.55) | (0.67-0.74) | (0.60-0.65) | (0.63-0.68) | (0.65-0.70) | (0.64-0.69) | **(0.71-0.76)** | (0.64-0.68) | (0.69-0.74) |
| 7-Year | 0.53 | 0.71 | 0.63 | 0.66 | 0.67 | 0.67 | **0.74** | 0.66 | 0.71 |
| AUC | (0.52-0.54) | (0.68-0.75) | (0.60-0.65) | (0.63-0.68) | (0.65-0.70) | (0.64-0.69) | **(0.72-0.76)** | (0.64-0.68) | (0.69-0.73) |
| 8-Year | 0.53 | 0.72 | 0.62 | 0.65 | 0.68 | 0.67 | **0.73** | 0.67 | 0.70 |
| AUC | (0.51-0.54) | (0.69-0.75) | (0.60-0.64) | (0.62-0.67) | (0.65-0.70) | (0.65-0.69) | **(0.71-0.75)** | (0.64-0.69) | (0.68-0.72) |
| 9-Year | 0.52 | 0.73 | 0.62 | 0.64 | 0.68 | 0.68 | **0.73** | 0.67 | 0.69 |
| AUC | (0.51-0.53) | (0.69-0.76) | (0.60-0.65) | (0.61-0.66) | (0.65-0.70) | (0.65-0.70) | **(0.70-0.75)** | (0.65-0.69) | (0.67-0.71) |
| 10-Year | 0.52 | 0.74 | 0.63 | 0.63 | 0.68 | 0.70 | **0.73** | 0.68 | 0.69 |
| AUC | (0.51-0.53) | (0.70-0.77) | (0.60-0.66) | (0.60-0.66) | (0.66-0.71) | (0.67-0.72) | **(0.71-0.75)** | (0.66-0.70) | (0.67-0.72) |
| Inhouse Normal BI-RADS Screening Test set: 5,139 exams, 102 followed by cancer diagnosis within 1 years; 404 diagnosis within 5 years; 612 diagnosis within 10 years. |  |  |  |  |  |  |  |  |  |
| 5-Year | 0.51 | 0.68 | 0.61 | 0.62 | 0.62 | 0.64 | **0.73** | 0.65 | 0.73 |
| C-index | (0.50-0.51) | (0.64-0.72) | (0.59-0.64) | (0.60-0.65) | (0.59-0.64) | (0.62-0.67) | **(0.71-0.75)** | (0.63-0.68) | (0.71-0.75) |
| 10-Year | 0.50 | 0.69 | 0.61 | 0.62 | 0.62 | 0.64 | **0.73** | 0.65 | 0.73 |
| C-index | (0.50-0.51) | (0.66-0.72) | (0.59-0.64) | (0.60-0.64) | (0.60-0.65) | (0.62-0.67) | **(0.71-0.75)** | (0.63-0.67) | (0.71-0.75) |
| 1-Year | 0.53 | 0.70 | 0.61 | 0.62 | 0.63 | 0.66 | **0.74** | 0.68 | 0.77 |
| AUC | (0.50-0.55) | (0.63-0.77) | (0.54-0.67) | (0.57-0.68) | (0.57-0.68) | (0.60-0.71) | **(0.69-0.79)** | (0.63-0.73) | (0.72-0.81) |
| 2-Year | 0.51 | 0.69 | 0.60 | 0.62 | 0.61 | 0.64 | **0.73** | 0.67 | 0.75 |
| AUC | (0.50-0.53) | (0.63-0.75) | (0.56-0.65) | (0.58-0.66) | (0.57-0.65) | (0.60-0.68) | **(0.69-0.76)** | (0.63-0.71) | (0.72-0.79) |
| 3-Year | 0.51 | 0.70 | 0.62 | 0.65 | 0.63 | 0.64 | **0.73** | 0.66 | 0.74 |
| AUC | (0.50-0.52) | (0.64-0.74) | (0.58-0.66) | (0.61-0.68) | (0.59-0.66) | (0.61-0.68) | **(0.70-0.76)** | (0.62-0.69) | (0.71-0.77) |
| 4-Year | 0.51 | 0.69 | 0.63 | 0.64 | 0.63 | 0.64 | **0.72** | 0.66 | 0.73 |
| AUC | (0.50-0.51) | (0.65-0.74) | (0.60-0.66) | (0.61-0.67) | (0.60-0.66) | (0.62-0.67) | **(0.70-0.75)** | (0.63-0.68) | (0.70-0.75) |
| 5-Year | 0.51 | 0.70 | 0.62 | 0.65 | 0.64 | 0.65 | **0.72** | 0.66 | 0.72 |
| AUC | (0.50-0.51) | (0.65-0.74) | (0.59-0.65) | (0.62-0.68) | (0.61-0.67) | (0.62-0.67) | **(0.70-0.75)** | (0.63-0.68) | (0.69-0.74) |
| 6-Year | 0.50 | 0.71 | 0.63 | 0.66 | 0.66 | 0.65 | **0.73** | 0.66 | 0.72 |
| AUC | (0.50-0.51) | (0.66-0.75) | (0.60-0.65) | (0.63-0.68) | (0.63-0.68) | (0.63-0.68) | **(0.71-0.76)** | (0.63-0.68) | (0.70-0.74) |
| 7-Year | 0.50 | 0.71 | 0.63 | 0.66 | 0.66 | 0.66 | **0.73** | 0.66 | 0.71 |
| AUC | (0.50-0.51) | (0.67-0.75) | (0.60-0.66) | (0.63-0.69) | (0.64-0.69) | (0.63-0.68) | **(0.71-0.76)** | (0.64-0.68) | (0.69-0.73) |
| 8-Year | 0.50 | 0.72 | 0.63 | 0.66 | 0.67 | 0.67 | **0.73** | 0.67 | 0.71 |
| AUC | (0.50-0.51) | (0.68-0.76) | (0.60-0.65) | (0.63-0.68) | (0.65-0.70) | (0.64-0.69) | **(0.71-0.75)** | (0.64-0.69) | (0.68-0.73) |
| 9-Year | 0.50 | 0.73 | 0.63 | 0.65 | 0.67 | 0.67 | **0.73** | 0.67 | 0.70 |
| AUC | (0.50-0.51) | (0.69-0.76) | (0.60-0.66) | (0.62-0.68) | (0.64-0.70) | (0.65-0.70) | **(0.70-0.75)** | (0.65-0.69) | (0.67-0.72) |
| 10-Year | 0.50 | 0.74 | 0.63 | 0.64 | 0.67 | 0.69 | **0.73** | 0.68 | 0.70 |
| AUC | (0.50-0.51) | (0.70-0.77) | (0.60-0.66) | (0.61-0.67) | (0.65-0.70) | (0.67-0.72) | **(0.71-0.76)** | (0.66-0.70) | (0.67-0.72) |

#### In normal BI-RADS screening population

In the normal BI-RADS screening test set with 5,139 examinations / 404 positives within 5 years / 612 positives within 10 years, we investigate the potential of risk models when radiologists cannot find any suspicious findings on the mammograms. Thus, the C-indices and AUCs of the BI-RADS are close to 0.5, as shown in Table 3 and Table S2. Our MTP-BCR with risk factors is still significantly better than all other methods among the extremely short-term with a 1-year AUC of 0.74 (95% CI, 0.69-0.79), long-term with a 5-year AUC of 0.72 (95% CI, 0.70-0.75), and longer-term with 10-year AUC of 0.73 (95% CI, 0.71-0.76) risk predictions. Especially in the BCSC model target population (aged 35-74, without prior BC history), the 1-year AUC of our MTP-BCR risk-based model reaches 0.85 (95% CI, 0.78-0.91) while the radiologists and the cancer detection-based models fail to outperform random guessing.

Consistent with the findings on the full inhouse test set, AUC and C-index metrics show that our MTP-BCR model performs similarly for unilateral BC risk prediction and patient-level cancer risk prediction on both screening sets. We note that using the full training dataset to finetune MIRAI lead to poor performance of MIRAI on these two screening sets (shown in Table S8 and S9, Replicate 5-Year BC Risk Prediction). Thus, we clean the training and validation sets using the same settings as for the two screening sets, and then re-finetune the MIRAI model and test it on the corresponding test set. Despite that, we find that the finetuned MIRAI model on the dataset with missing screening follow-up seems to be difficult. Specifically, the performances of the finetuned MIRAI from 3- to 10-year BC risk prediction become worse (shown in Fig. 3 B and C). Thus, for a meaningful comparison, it is necessary to re-implement these comparison experiments on the two recollected five-year screening datasets which cleaned exams with missing follow-up labels. As shown in Table S8 and S9, the AUCs and C-indices of MIRAI, after finetuning, reaches its own optimal, and outperforms the 5-year BCSC model. The finetuned MIRAI only achieves similar performance to our image-only MTP-BCR model from 3- to 5-year risk at both screening test sets (when only for women who can be scored by the BCSC model) (Table S9). Yet our method with risk factors still surpasses the MIRAI.

### The ability of short and long future BC risk assessment

While the above results demonstrate the advantages of our methods in risk prediction, the ability to predict real long-term future BC risk after eliminating the biases of cancer detection and short-term to 5-year risk prediction from current mammograms requires further exploration. Thus, we compare the models’ performance in 5-year and 10-year risk prediction in the different subgroups of the full inhouse test sets by excluding exams from women diagnosed with cancer within less than 1, 3, and 5 years. These results, as shown in Table 4, demonstrate that our methods could not only detect BC and improve the performance of 5-year risk prediction compared with other SOTA methods but also learn the features related to the real longer-term (10 years) risk. The results of replicate 5-year BC risk prediction are shown in Table S10.

**Table 4.** Comparison of future risk predictions. AUC results are presented with 95% Confidence Interval. BCSC: Breast Cancer Surveillance Consortium; GMIC: Globally-Aware Multiple Instance Classifier. Note that results of the BCSC model are based on part of the full inhouse test set, as they are out of the age range of 35-74 or with prior BC history. For fair comparison we also impended the comparison experiments excluding the women that did not score across the BCSC model. Bold: *P* < 0.05, the AUCs of our methods are significantly higher than all other model for the same time horizon.

| Use Risk Factors | BI-RADS | BCSC | GMIC | MIRAI Test | MIRAI Finetune | MTP-BCR (Ours) Patient Level |  | MTP-BCR (Ours) Unilateral Breast Level |  |
| --- | --- | --- | --- | --- | --- | --- | --- | --- | --- |
|  | - | Yes | No | - | - | No | Yes | No | Yes |
| **Full test set** |  |  |  |  |  |  |  |  |  |
| 2-5 Year | 0.51 | - | 0.63 | 0.64 | 0.60 | 0.64 | **0.72** | 0.65 | 0.71 |
| AUC | (0.50-0.52) | - | (0.60-0.66) | (0.61-0.67) | (0.57-0.63) | (0.61-0.66) | **(0.69-0.74)** | (0.62-0.67) | (0.68-0.73) |
| 4-5 Year | 0.50 | - | 0.61 | 0.62 | 0.58 | 0.63 | **0.72** | 0.63 | 0.69 |
| AUC | (0.49-0.52) | - | (0.57-0.65) | (0.58-0.66) | (0.53-0.63) | (0.59-0.66) | **(0.68-0.75)** | (0.59-0.67) | (0.66-0.73) |
| 2-10 Year | 0.50 | - | 0.63 | 0.62 | 0.60 | **0.68** | **0.73** | 0.67 | 0.69 |
| AUC | (0.49-0.51) | - | (0.60-0.65) | (0.59-0.65) | (0.57-0.62) | **(0.66-0.71)** | **(0.70-0.75)** | (0.65-0.69) | (0.66-0.71) |
| 4-10 Year | 0.50 | - | 0.61 | 0.60 | 0.59 | **0.67** | **0.73** | 0.66 | 0.68 |
| AUC | (0.49-0.51) | - | (0.58-0.64) | (0.57-0.63) | (0.56-0.62) | **(0.64-0.70)** | **(0.70-0.75)** | (0.64-0.68) | (0.66-0.71) |
| 6-10 Year | 0.50 | - | 0.60 | 0.58 | 0.60 | **0.67** | **0.73** | 0.65 | 0.69 |
| AUC | (0.49-0.52) | - | (0.57-0.64) | (0.55-0.62) | (0.56-0.63) | **(0.63-0.70)** | **(0.70-0.76)** | (0.62-0.68) | (0.66-0.72) |
| **Part of Full test set: only women who completed scored across the BCSC model** |  |  |  |  |  |  |  |  |  |
| 2-5 Year | 0.52 | 0.69 | 0.66 | 0.67 | 0.59 | 0.69 | **0.76** | 0.69 | 0.74 |
| AUC | (0.49-0.55) | (0.64-0.74) | (0.61-0.71) | (0.62-0.71) | (0.54-0.65) | (0.64-0.74) | **(0.72-0.80)** | (0.65-0.74) | (0.70-0.78) |
| 4-5 Year | 0.50 | 0.67 | 0.64 | 0.64 | 0.57 | 0.68 | **0.76** | 0.66 | 0.73 |
| AUC | (0.47-0.53) | (0.60-0.74) | (0.58-0.71) | (0.57-0.70) | (0.49-0.64) | (0.61-0.74) | **(0.71-0.81)** | (0.60-0.73) | (0.68-0.78) |
| 2-10 Year | 0.52 | 0.73 | 0.63 | 0.64 | 0.57 | 0.73 | **0.79** | 0.72 | 0.72 |
| AUC | (0.50-0.54) | (0.69-0.77) | (0.59-0.67) | (0.60-0.69) | (0.53-0.62) | (0.69-0.77) | **(0.75-0.82)** | (0.68-0.75) | (0.68-0.75) |
| 4-10 Year | 0.51 | 0.72 | 0.62 | 0.62 | 0.56 | 0.72 | **0.79** | 0.70 | 0.72 |
| AUC | (0.49-0.53) | (0.67-0.76) | (0.57-0.67) | (0.57-0.67) | (0.51-0.61) | (0.68-0.76) | **(0.75-0.82)** | (0.66-0.74) | (0.68-0.75) |
| 6-10 Year | 0.51 | 0.71 | 0.61 | 0.61 | 0.56 | 0.70 | **0.79** | 0.68 | 0.73 |
| AUC | (0.49-0.54) | (0.66-0.76) | (0.55-0.67) | (0.55-0.66) | (0.50-0.63) | (0.65-0.75) | **(0.75-0.83)** | (0.64-0.73) | (0.68-0.77) |

### Clinical sub-group analysis

To distinguish how our MTP-BCR model performs in different populations and to determine the potential population that can benefit most from it, we evaluate our risk model in different clinical subgroup, based on age, breast density, personal history, and future cancer sub-types (Fig. 4). We find that the MTP-BCR model performed similarly across different density groups and independent from future cancer sub-types. The C-indices for the MTP-BCR model for women aged <40, 40-60, and 60-80 are 0.87 (95% CI, 0.84-0.90), 0.81 (95% CI, 0.79-0.83) and 0.82 (95% CI, 0.79-0.84), respectively, which implies that our risk model performs better in younger cohorts. We also note that the risk model performs best in the female population without any prior personal BC history with C-index of 0.86 (95% CI, 0.84-0.87), compared to 0.76 (95% CI, 0.74-0.78), for women with a personal history of breast cancer. We also compare the C-index of 10-year risk prediction of different methods in different sub-groups, which are available in Table S3. Our results are further supported by the consistent performance from additional subgroup analyses in recollected 5-year risk dataset (Tables S11, supplemental). Due to a lack of race labels, the group analysis for different race groups is not performed.

**Fig. 4.**
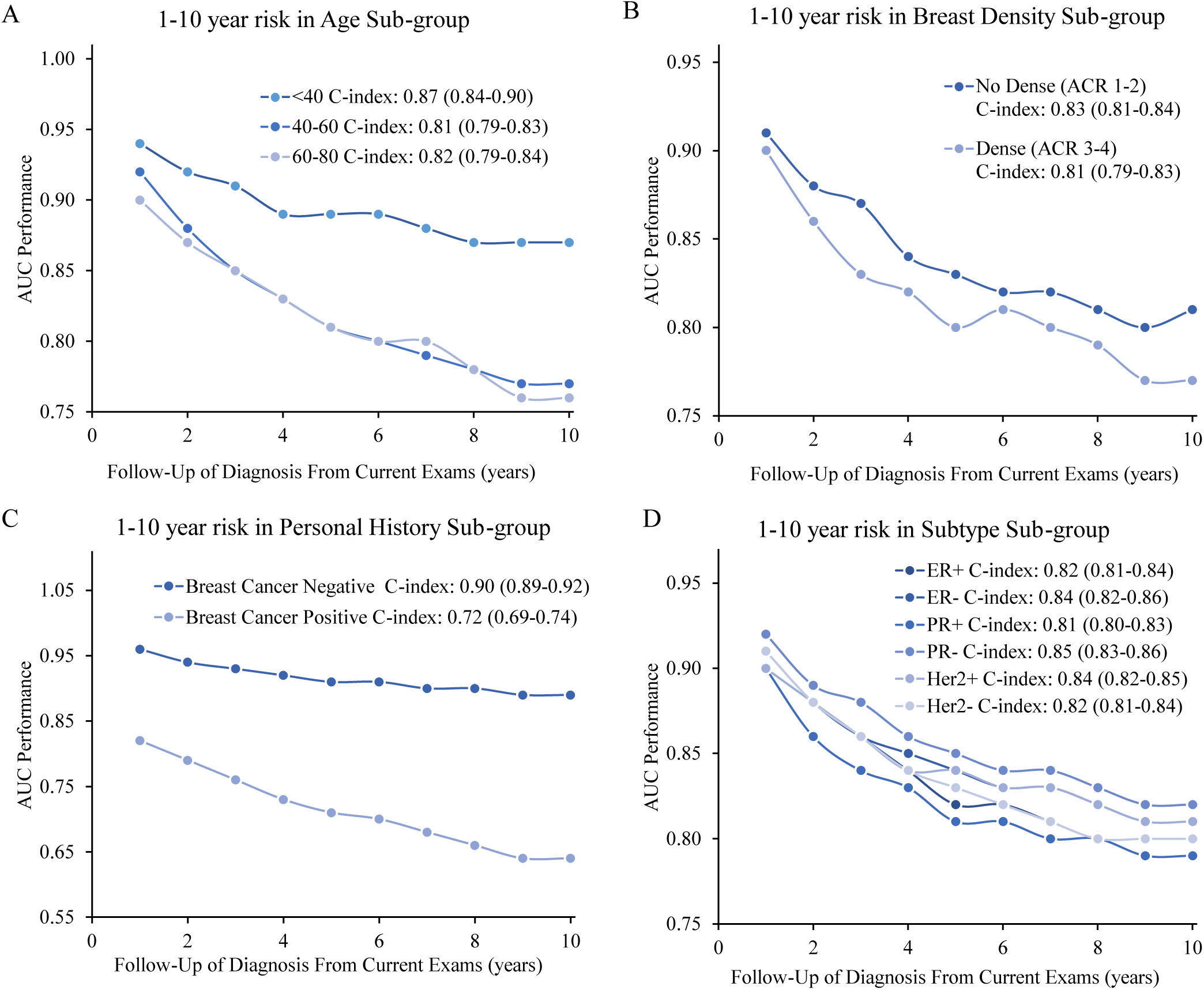
Cumulative risk at multiple time points on different sub-groups.

### Consistency of model attention in longitudinal images

To investigate how risk-related areas evolve on multiple-time mammograms that our MTP-BCR model focused on, we utilize the gradient-weighted class activation maps (Grad-CAM) (*36*). Fig. 5 shows a visualization example of a BC patient. The heatmaps highlight potentially related regions where our proposed MTP-BCR model identifies predictive imaging features for BC risk. While this visualization is a preliminary process, results show that the high-risk regions from multiple time point examinations that our model focuses on are relatively consistent. Moreover, the heatmaps show that our risk model could accurately figure out high-risk areas of short-term BC at both the craniocaudal (CC) and the mediolateral oblique (MLO) views of mammograms. The similar result is observed from the heatmaps of the retrained model on the inhouse five-year risk dataset (Fig. S5, supplemental).

**Fig. 5.**
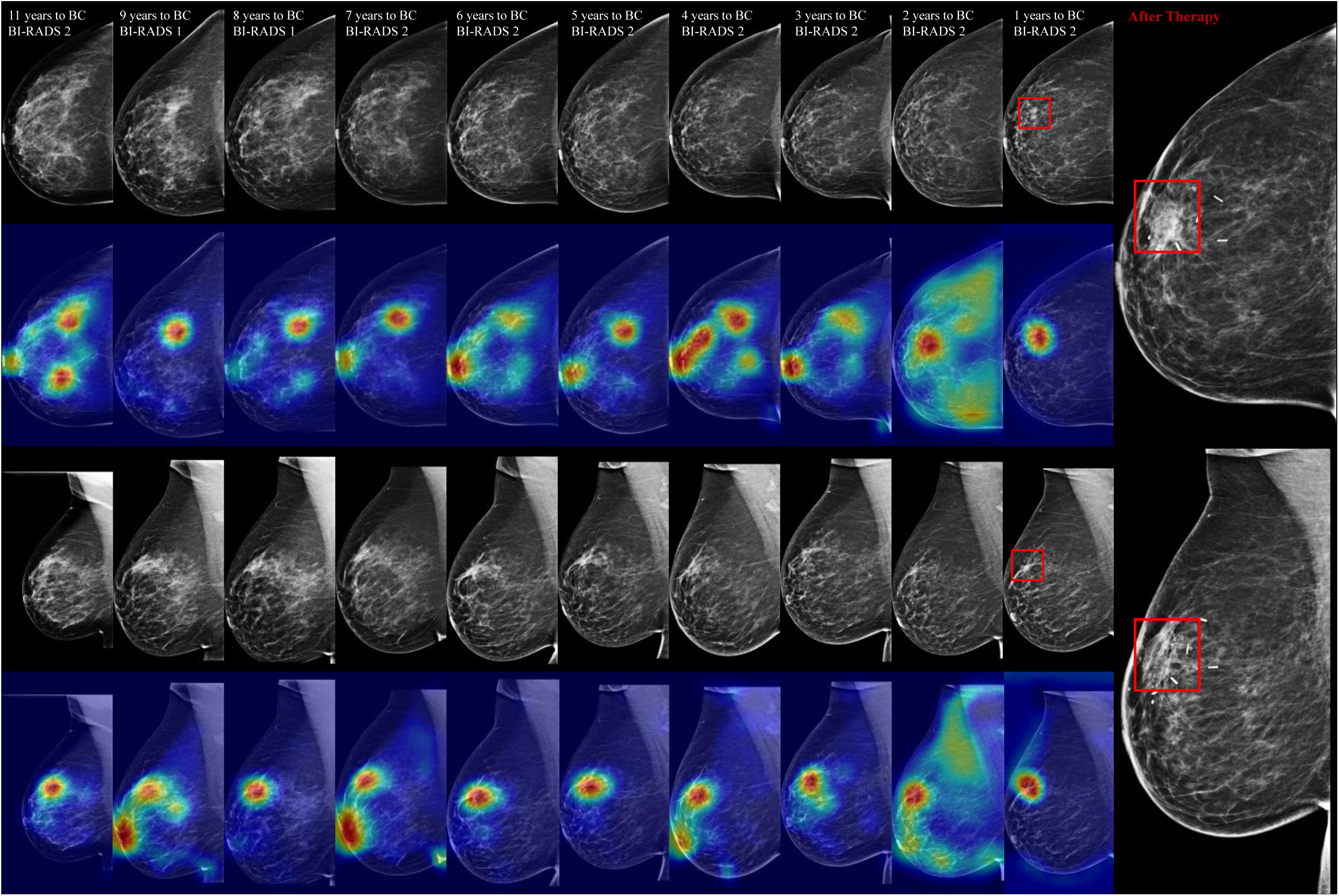
An example of class activation map (CAM) visualization. The longitudinal craniocaudal (CC) and mediolateral oblique (MLO) mammograms were acquired from a patient who participated in ten consecutive breast cancer screening from 2005 to 2015, culminating in a breast cancer diagnosis during the last screening (invasive ductal and lobular carcinoma located at C50.4, exhibiting positive expression of estrogen receptor (ER+), progesterone receptor (PR+), and human epidermal growth factor receptor 2 (Her2Neu+)). The closer to red, the more relevant the pixel is to the risk prediction.

## Discussion

In this study, we develop a multi-time-point examination-based risk model, MTP-BCR, to assess 10-year breast cancer risk on patient and unilateral breast level using longitudinal screening mammograms and medical records. From extremely short-term risk (1-year, BC detection) to long-term (10-year) risk prediction, MTP-BCR outperforms radiologists’ BI-RADS assessment, a SOTA BC detection method (GMIC), an extensively clinically validated single time point-based 5-year risk DL approach (MIRAI), and a traditional clinical risk model (BCSC). Apart from the patient level risk, our method could also estimate the risk base on unilateral breast level with comparable ability. Experiments on different screening subcohorts suggest that the longitudinal assessment of MTP-BCR is able to accurately identify longer-term, future risk-related features on mammograms, which is further supported by consistent heatmaps of multi-time point mammograms. Final sub-group analysis indicates that the proposed method performs consistently across subgroups of different breast densities and for future types of BC.

The motivation to develop BC risk models is for guiding personalized screening or triggering prevention regimens. The idea is to determine the screening frequency and the appropriate screening modality based on the individual risk of women, potentially also to recommend preventive therapy for women at a high-risk of developing breast cancer (*13*). Based on of risk factors such as age, genetic determinants, family history, previous benign biopsies, and recently considered image-based breast density, traditional risk models are used to globally assess 5-year, 10-year, or lifetime risk for large groups of women (*37*). However, our results show that density alone is not sufficient to represent all of the risk-related information within the mammograms. Moreover, ignoring the short-term risk of BC limits the value of these models in early BC detection. Recent DL-based risk models may fully utilize screening images but mainly target short-term risk prediction or interval cancer detection while ignoring long-term risk, which limits the ability to offer personalized screening recommendations and preventive interventions. In contrast our MTP-BCR risk model combines the advantages of both short- and long-term risk prediction strategies.

For short-term risk estimation, our risk model outperforms radiologists’ BI-RADS and other recent DL methods. The results on the full test set demonstrate that our MTP-BCR risk model is more competitive than other risk models in indicating current or future BC risk in a realistic complex clinical screening setting independent of radiological interpretation. Other methods, especially traditional risk tools, can only work with modest performance to estimate women’s future risk on the premise that the physician confirm that there is no existing cancer. Furthermore, in the normal BI-RADS screening group, our risk model has the highest AUC of 0.74 for the 1-year risk prediction, which can be regarded as an aid for radiologists to improve interval cancer detection in the whole screening population, including primary and recurring cancers. When detecting the primary interval cancers of the population that the BCSC model targets, our risk model reaches an AUC of 0.87. Aiming to facilitate radiologists’ understanding of the model decision-making, our risk model could also estimate the unilateral-breast level risk with similar performance to the patient-level risk prediction. Thus, our retrospective analysis and visualizations indicate that the MTP-BCR risk model could improve early detection and reduce interval cancer by guiding radiologists to identify high-risk regions on images.

For 5-year risk prediction, our methods surpass the SOTA MIRAI risk method across all screening cohorts or subcohorts. Although the MIRAI model does not need to have the risk factors as inputs and performs similarly to our image-only MTP-BCR model at 5-year risk (*P* > 0.05) on the biopsy negative screening set and normal BI-RADS set, we note that the MIRAI model involves a pretrained risk factors predictor enabling it to benefit from the missing risk factors that reaches a similar performance of 5-year risk prediction as MIRAI with risk factors (*P* = 0.27) (*19*). While our MTP-BCR model with risk factors is significantly better than all others (All *P* values < 0.05). Besides, the MIRAI is already trained on their large private MGH dataset. The training set includes 210,819 exams is 6.5 times larger than our Inhouse training set. Then we still fully finetuned the MIRAI with its weight in our In-house training set without freezing the encoder. On the other hand, our MTP-BCR model is only trained on the In-house dataset directly. As for long-term risk, the MTP-BCR risk model is more accurate than the BCSC 10-year risk model, which suggests our risk model has the potential for better decisions regarding a risk-adapted screening regimen and preventive therapy. At breast level, risk could be the foundation of more refined screening and prevention strategies. It should be underlined that in this study we also involve patients with prior BC history, which are not included in most of the risk models. Our risk model is, in fact, also designed to leverage the history information of prior-tumor and therapy, which the recurrence risk models use (*38*), for recurrence cancer risk prediction.

The promising performance of MTP-BCR can be attributed to its capacity to capture unique BC risk-related characteristics. The multi-task learning strategy helps the model to fully extract risk-related features from images and also improves the generalization of the DL model (*39*). The multi-level learning strategy enables our model to learn the relationship between local (unilateral breast) level risk and global (patient) level risk while keeping the local information as much as possible when combining the local features for the summary of the global features. Thus, the MTP-BCR risk model can consistently focus on similar regions on longitudinal mammograms without registration. For accurate longer-term BC risk prediction, we need not only the static risk-related features from the single time point exam but also the dynamic features from the multi-time point exams to indicate the development risk of BC. Recently, a study (*20*) also explored the potential of using longitudinal mammogram examinations to improve short-term risk prediction. But we note that it has been restricted to a small image-only dataset and a setting of a fixed number of input time points, which hinders its clinical application. On the contrary, our MTP-BCR model is built based on an extensive clinical screening mammogram dataset, enabling efficient use of risk factors, and has the flexibility to input 0-5 historical reference exams.

This research has limitations. Although we leverage the prior tumor information and therapy records (as explained in Section Methods) to improve our risk model’s performance on BC recurrence risk, big gaps exist between the subgroups with/without prior BC. More efforts are needed to improve the performance of recurrence risk prediction. Further validation of our model is required before it can be broadly implemented in clinical practice. For instance, more detailed demographics (e.g., race) are required to prove its generalizability. We note that the breast cancer incidence in our screening set is higher than that of the standard screening dataset. Therefore, in the future, we will also explore the clinical potential of our MTP-BCR model based on external standard screening mammography datasets from multiple hospitals. To validate the detection of extremely early signs of BC, a pixel-level annotated dataset is necessary. Moreover, a reader study for incorporation of the risk model in the radiologist workflow may also be a future direction for further demonstrating the benefits of risk models for personalization BC screening policy.

In conclusion, we propose a novel DL model using longitudinal mammogram examinations and history obtained from medical records that outperforms the SOTA MIRAI and traditional BCSC risk model by a large margin. The improvement is consistent across screening and future risk subgroups. These results support the hypothesis that longitudinal mammography contains informative spatiotemporal indicators of future breast risk that cannot be captured by the single-time point DL models. Multi-time point models based on longitudinal analysis strategies have the potential to replace single-time point based risk prediction models. Apart from increasing the accuracy for BC risk prediction, we also improve the interpretability of our risk model, which could potentially accelerate the translation of personalized AI-based risk stratification into routine BC screening policies.

## Materials and Methods

### Data collection

Our retrospective study was approved by the Institutional Review Board (IRB) of Netherlands Cancer Institute (NKI) with protocol numbers: IRBd21-060. A flowchart illustrating the construction of this large study dataset is shown in Fig. 2. We collect 37,517 patients recorded in our hospital between January 1, 2004, and December 31, 2020. Then we collect the longitudinal digital screening mammograms and exclude patients without at least one year of screening follow-up, in line with the research (*19*). Details about the distributions of the dataset are available in Fig. S1. Although part of the patients did not have 10-year screening follow-up, we also leverage their known outcomes and images to supervise the model. Therefore, we keep 9,133 patients consisting of 2,562 BC patients who were biopsy-proven within 10 years and 6,571 at intermediate risk who had at least 10 years of screening follow-up and did not receive a cancer diagnosis. All patients are randomly divided into training, validation, and test sets with a ratio of 7.5:1:1.5. The training, validation, and test sets include 6,858, 919, and 1,356 patients with 32,049, 4,432, and 6,311 examinations, respectively.

BC-relevant risk factors are already showing an essential role in both traditional (*12, 15*) and image-based DL methods (*16, 19*). In our study, we collect the risk factors through electronic medical records from clinical radiology, tumor, therapy, and pathology reports from our hospital. The distribution of clinical risk factors in the inhouse dataset is shown in Table 1. Specifically, we obtain age, race, BI-RADS and breast density (ACR) grade, family history, genetic determinants, previous BC history, previous ovarian cancer history, self-reported menopausal status, and age of menarche. The BI-RADS and breast density (ACR) grade are estimated by radiologists during clinical interpretation. BI-RADS grades include additional imaging required (BI-RADS 0), normal (BI-RADS 1), benign (BI-RADS 2), probably benign (BI-RADS 3), suspicious for malignancy (BI-RADS 4), highly suggestive of malignancy (BI-RADS 5), and known biopsy-proven malignancy (BI-RADS 6). ACR class include mostly composed of fatty tissue (ACR 1), scattered fibroglandular tissue (ACR 2), heterogeneously dense (ACR 3), and extremely dense (ACR 4). Images with missing densities are interpolated by nearest neighbour interpolation with reference to the density estimates of screening images from adjacent years of the patient. Because the weights of patients are missing, we did not calculate the body mass index (BMI). In our study, we also include patients with prior BC. Thus, following the research of recurrence risk prediction (*38*), we also leverage the information of prior tumor, which include pathologic tumor (pT)-stage, pathologic node (pN)-stage, hormone receptor status (estrogen receptor (ER)- and progesterone receptor (PR)-status), anti-hormonal therapy, human epidermal growth factor receptor 2 (HER2-status), type of surgery, adjuvant chemotherapy, adjuvant radiation therapy, antibody therapy and Pathologic Complete Response (pCR).

### Problem formulation

For the risk prediction, we first divide the relevant time span into one-year time-slots and treat each slot as an independent class. To evaluate the overall risk of the first *j* years, the probabilities of each year are summed together from the first year up to the *j*_*th*_ year. The formulas are defined as follows Eq. 1:

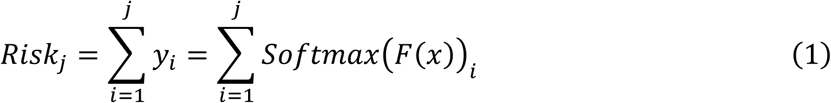

where, *y*_*i*_ means the predicted probability of an exam getting BC diagnosis at *i*_*th*_ year, which is calculated by inputting a sequence of a mammograms and corrected risk factors *m* into the model *F* and using the *Softmax* function for the probability generalization. For example, as shown in Fig. 1B, to predict the *Risk* of a patient getting BC within *j* = 2 years from the checked images, it can be calculated as the sum of the probability of the first year and second year.

### Architectural details

As shown in Fig. 1, the MTP-BCR model consists of the network weights shared **Image Encoder (*φ*_*encoder*_)** connected by **Side-Specific (Unilateral-based) Module (*φ*_*unilateral*)_**, **Exam-based Module (*φ_exam_*)**, and finally, **Multi-Time-Point fusion Module (*φ_fusion_*)** combines with the inputted risk factors of patients. Moreover, to improve risk modeling performance and generalization, we also introduce **multi-task learning**, which could benefit from learning the domain-specific features from multiple BC risk-related tasks (detailed below).

#### Image Encoder

We employ ImageNet pre-trained ResNet-18, excluding the last full connection (FC), as the encoder (*φ_encoder_*) to extract breast tissue features. The weights-shared encoders correspond to each image from a sequence of mammography exams. Each exam includes four images including craniocaudal (CC) view (*v* = *cc*) and mediolateral oblique (MLO) view (*v* = *mlo*) from the left(*l* = *left*) and right(*l* = *right*) side breast. Thus, shown in the Eq. 2, each input mammogram, 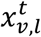, from the 6 time point (*t*) exams is represented as the high-dimensional locally feature vector 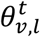 with the size of 512 × 1 by the encoder separately.

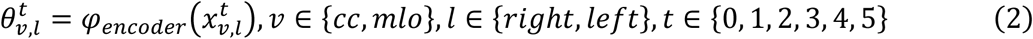

#### Side-Specific Prediction Module

Ipsilateral CC and MLO views are different projection views of the same breast. Practically, they are combined to express the three-dimensional structure of the breast, which radiologists use to detect abnormalities. Moreover, most tumors only appear in one of the breasts (*40*). In order to train the model to learn the three-dimensional structure of the breast fully and correspond to the previous left and right breast-specific tumor information, we concatenate (⊕) the feature vectors of the ipsilateral view, combined with the side-specifically prior tumor information 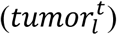 and then place a Multi-layer Perceptron (MLP) for side-based multi-task learning. The MLP layer includes two FC layers with an input size of 1152 for the first FC layer and 512 output units each. A dropout layer with a rate of 0.5 between the two FC layers. Therefore, show in Eq. 3 features of the ipsilateral CC and MLO views and outputs a vector 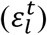 with the size of 512 × 1 representing the unilateral-based breast features.

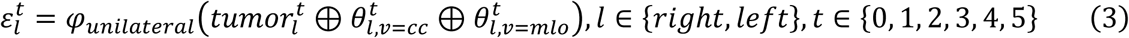

#### Exam-Based Prediction Module

For a similar purpose, we also need to combine the information of bilateral breasts to predict the patient level risk. As in Eq. 4, we concatenate the feature vectors from the output bilateral breasts and feed them to another MLP layer with the same structure for the exam-based multi-task learning. Also, a size of 512 × 1 vector feature (*δ*^*t*^), which combines the features from right and left breast, represents the global information of a four-view exam.

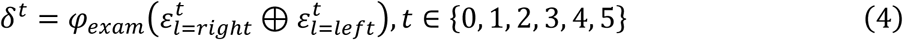

#### Multi-Time Point Fusion Model

For learning the risk development pattern from the longitudinal screening mammograms, five historic exams before the current exam are randomly selected as the reference for the comparison by the multi-time point fusion model. The current exam refers to the target exam for which we access future BC risk. A sequence of mammography exams serve as references along with the time intervals (*i*_0_, *i*_1_, …, *i*_5_) to the current exam and are combined with the risk factors to predict the future likelihood of BC occurring after the current mammograms. Inspired by the research (*26*), we leverage a sequence/time-aware transformer learning (*41*) to capture features about the temporal relations between multiple mammograms, which aims to disentangle the risk-relevant changing patterns from the normal breast tissue changing patterns. For embedding the spatiotemporal relationships of past-current exams into the continuous latent space, we employ Continuous Position Embedding (CPE) method (*26*), which computes time continuous embedding *e*^*t*^ to condition the image features. Not that, to avoid ignoring local information of images during multi-time exam comparison, we combine both the local image features 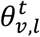, and global features *δ*^*t*^. For patients without five history records for references, we select all history records and then mute the missing data by filling 0. At the same time, for the purpose of data augmentation, we randomly drop a subset of exams in the reference sequence to improve model robustness and avoid overfitting. The fusion model also includes the patient risk factors (*riskf*). Subsequently, a fused feature (*τ*, a vector size of 640 × 1) is obtained for the final multi-time fused-based multi-task learning, representing the patient’s multi-time point screening information.

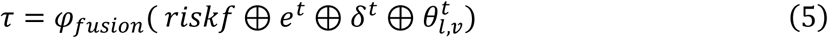

#### Multi-task classifier for side-based, exam-based, and multi-time fused prediction

During the multi-task learning, we constrain the feature extractor for BC risk-related prediction task learning, shown in Fig. 1C. For instance, the predictions of breast-based BC risk, history, tumor location, and tumor sub-type are included for the side-based multi-task classifier. For the exam-based multi-task classifier, we replace the breast-based BC risk prediction and history prediction with exam-based prediction. We also add the prediction of age, BI-RADS, race, density, and manufacture. And for the final multi-time fused classifier, we mainly focus on unilateral specific BC risk. We calculate the Binary Cross Entropy (BCE) for risk prediction and Cross Entropy (CE) Loss for other predictions. For all three classifiers, we mainly focus on the task of risk prediction thus risk-specific tasks have 5 times higher weight than other tasks during the training. For total loss computing, Eq. 6, we also allocate weight *w*_*fusion*_ = 1 to the final multi-time point fused classifier, 5 times higher than the other two classifiers (*w*_*side*_ = 0.2, *w*_*exam*_ = 0.2). We choose the weights of loss after the hyperparameter search.

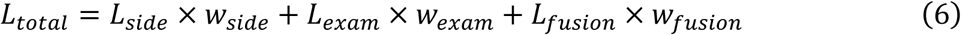

### Implementation details

We use ResNet-18 initializing with ImageNet pre-trained weights as the backbone of all our methods. All methods are implemented in PyTorch (version 1.12.1) with the same training strategies. We use the Adam (*42*) optimizer and a rate of 0.5 for dropout (*43*) after every fully connected layer. We train models for 20 epochs with a batch size of 8 and an initial learning rate of 10^-4^. The learning rate is decayed by a factor of 10 every five epochs. The best models of each method are chosen with the best AUC performance index on the validation set. The experiments are performed on a Quadro A6000 GPU (48GB). The source code is available at https://github.com/Netherlands-Cancer-Institute/MTP-BCR. Mammograms with standard DICOM format are pre-processed before being fed into the model. First, we convert the images into 16-bit PNG format and segment the whole breast region to exclude the background. Then, to unify the size of all images, we zero-pad and resize images to 512 by 1024 pixels while retaining the relative scale and aspect ratio. Finally, the image is normalized using the min-max method. We also employ standard data augmentation techniques (i.e., random flip, brightness, and contrast) during training for model robustness and overfitting prevention.

### Evaluation metrics and statistical analysis

In this study, the tasks of prediction of 1- to 10-year risk are categorical classification tasks, in which positive samples are the patients diagnosed with BC within 1 to 10 years while negative samples are women who stayed healthy for at least 1 to 10 year-screening follow-ups. The performances of the different methods are evaluated by the area under the receiver operating characteristic curve (AUC, calculated by scikit-learn, version: 1.1.2, https://scikit-learn.org). To generally evaluate AUCs across all times (from 1- to 10-year risk), the Uno’s C-index (*44*) is calculated using scikit-survival (version 0.18.0, https://scikit-survival.readthedocs.io/en/stable/). The 95% confidence intervals (CI) of AUC and C-index matrices are estimated by bootstrapping with 1,000 bootstraps for each measure. Statistical significance among different methods is assessed using DeLong’s test (*45*), with the significant level predefined as *P* < 0.05.

## Data Availability

The inhouse dataset is used under license to the respective hospital system for the current study and is not publicly available.

## Acknowledgments

The authors thank to the support from the Chinese Scholarship Council scholarship (CSC) (X.W., Y.G., and L.H.: 202107720016, 202006930001, and 202006240065) and Guangzhou Elite Project (T.Z.: TZ–JY201948).

## Author contributions

Conceptualization: X.W., T.T., and R.M.Data Collection: X.W., T.Z., L.H., and Y.G. Methodology: X.W., T.T., Y.G., R.M, and R.S. Investigation: X.W., Y.G., and A.D.A. Visualization: X.W., Y.G., and R.S. Supervision: T.T., R.M., and R.B.T. Writing—original draft: X.W., Y.G., R.S., T.Z., and L.H. Writing—review & editing: R.M., N.K., X.W., C.A.D., M.K.S., and J.T.

## Data and materials availability

All custom codes related to training and developing the deep learning models is available on https://github.com/Netherlands-Cancer-Institute/MTP-BCR. <u>The trained MTP-BCR model is available after the publication of this paper.</u> The inhouse datasets are used under license to the respective hospital system for the current study and are not publicly available. All data associated with this study are present in the paper or the Supplementary Materials.

